# Master protocols in Low-and-Middle income countries: A review of current use, limitations, and opportunities for precision medicine

**DOI:** 10.1101/2024.12.03.24318453

**Authors:** Luke Ouma, Sarah Al-Ashmori, Samuel Sarkodie, Lou Whitehead, Ann Breeze Konkoth, Shaun Hiu, Theophile Bigirumurame, Dorcas Kareithi, Jingky Lozano-Kuehne, Marzieh Shahmandi, James M.S. Wason

## Abstract

**Background:** Master protocols - umbrella, basket and platform trials that study multiple therapies, multiple diseases or both, offer many advantages, most profoundly that they answer multiple treatment related questions, that would otherwise take multiple trials. We conducted a review of trial registries to characterise their use in advancing precision medicine in low and middle income countries (LMICs).

**Methods:** We searched trial records available in 20 trial registries globally, including ClinicalTrials.gov and WHO ICTRP, to identify umbrella, basket and platform trials launched until 30 September 2023.

**Results:** We identified 102 master protocols - 29 umbrella trials, 31 basket trials, 36 platform trials, as well as 6 other designs that partially aligned with the working definition of master protocols run in 54 different LMICs. Most trials were pharmaceutical industry-sponsored studies (60/102, 58.8%), conducted in oncology settings (56/102, 54.9%), currently ongoing (69/102, 67.6%) in early phase (phase I and II) settings (70/102, 68.6%). There was a greater representation of upper middle-income countries, particularly China that was a site to more than half of all master protocols (53/102, 52%). Other common countries included Brazil, Russia, Turkey and Argentina. Most master protocols (93/102 91.2%) have been planned or launched in the last five years (2019 onwards), mainly with international collaborations in high-income countries. Only a small proportion of trials (5/102, 4.9%) launched exclusively in LMICs excluding china and European LMICs. For most studies, the statistical aspects of trial design and trial documentation (including study protocols and statistical analysis plans) were not publicly accessible.

**Conclusion:** Unlike high-income countries, where several hundreds of master protocols are ongoing or completed, there is limited use of master protocols in LMICs, partly owing to low penetration of precision medicine research and limited clinical trial infrastructure in most LMICs. The evidence presented herein create a case for supporting precision medicine initiatives in LMICs (especially Africa), and training and capacity building initiatives focused on innovative clinical trial designs like master protocols, especially in therapeutic areas outside oncology.

## 1 Introduction

Master protocols, comprising umbrella, basket and platform trials, have revolutionised the drug development landscape in many therapeutic areas by allowing the study of multiple therapies, multiple diseases or both under a single trial infrastructure [1]. Today, several disease settings, including COVID-19 [2], cancer [3, 4], immune-mediated inflammatory diseases [5]and Alzheimer’s disease [6], have all benefited from efficient drug trials, faster development of interventions, multiple promising targeted therapies and improved patient outcomes overall, thanks to master protocol trial designs.

Today, many reviews have characterised the advantages of master protocols in terms of operational efficiency and logistical advantages, including faster recruitment given a centralised screening and assaying platform, standardisation across sub-studies, and simultaneous approval of sub-studies. Platform trials adaptively allow therapies to enter and leave the trial. During the COVID-19 pandemic, these designs were very popular [7] [8] [9], facilitating the timely investigation of many therapies - in the RECOVERY trial [9], decisions on at least 10 experimental therapies were conclusive in under two years. Basket trials have spearheaded the precision medicine agenda by evaluating single or combination therapies in several diseases that share a common characteristic [1], thus enabling trials of therapies even in small disease populations. For example, in 2006, imatinib was approved by the U.S. Food and Drug Administration for five different types of rare and life-threatening cancers on the basis of a phase II single-arm basket trial [10]. Another type of master protocol design, Umbrella trials, have enabled the concurrent investigation and development of various targeted therapies within a single disease setting [11], leading to the approval of biomarker-targeted therapies such as erlotinib and crizotinib in non small-cell lung cancer [12].

An examination of the geographic distribution of master protocol use, reveals that the rise in master protocol use in the last decade is mostly confined to high-income countries, usually involving early phase trials in oncology. Park and colleagues [13] in their 2019 review reported that only 6/83 master protocols (VE-BASKET [14], FUTURE [15], TRUMP [16], EBOLA [17], GBM AGILE [18], DIAN-TU [6]) had an LMIC country representation. Together, these 6 studies spanned only three LMICs: China, Brazil, and Mexico. In other systematic reviews of master protocols [19, 20], the geographic coverage of master protocols by country was not characterised.

Furthermore, the quantity of clinical trial research in LMICs remains inadequate, despite experiencing the greatest burden of disease globally [21, 22]. The greater disease burden in LMICs, rapidly changing healthcare needs, need for timely evaluation of promising (drug and non-drug) interventions (as in the EBOLA or COVID-19 pandemic), arguably justify the need to design and deliver innovative clinical trials across a broad range of diseases and treatment types in LMICs. Currently, the rarity of master protocols in LMICs can be attributed in part to the focus on published studies and limited awareness of their utility beyond oncology and biomarker-driven studies. Besides, master protocols can face greater operational and regulatory challenges [23] especially in regions with limited trial infrastructure such as LMICs, thus limiting their use to settings such as China and European middle-income countries where trial infrastructure is fairly well-developed.

Here, we present a review of clinical trial registries that seeks to unravel a clearer representation of the current landscape, including all studies that have been planned, launched and/or stopped early. Through a clearer understanding of the *status quo*, we develop a case for improving the use of master protocols in LMICs across various diseases, reflecting on some concepts, potential barriers, opportunities for their use, and a way forward.

## 2 Methods

### 2.1 Data sources and search strategy

We conducted a search of registered clinical trials present in 20 different trial registries across the world, that provide information to the World Health Organisation (WHO) International Clinical Trials Registry Platform (ICTRP) [24]. Our search included trial records registered since 01 July 2005 (enforcement of mandatory trial registration by ICJME) up to 30^th^ September 2023. A full list of these registries is provided in supplementary material section 2 including Clinicaltrials.gov [25], Pan African Clinical Trials Registry [26] and EU clinical trials register [27]. A comprehensive search (see supplementary material) was run in clinicaltrials.gov [25] and WHO ICTRP. In addition, we conducted manual forward/backward citation checks of relevant trials from recent reviews of master protocols.

### 2.2 Inclusion criteria

We included registered trials conducted in at least one or more sites in a LMIC country, according to the World Bank income classification of countries [28]. Given the common mislabelling of the different master protocols in the literature, we adopted the standardised definitions in Table 1, and re-categorised the trials if they were misclassified according to our working definitions. When a master protocol did not neatly fit the definitions of either umbrella, basket or platform trial - by combining elements of more than one master protocol - it was included and categorised as a complex design. All trial records were independently assessed for inclusion by two reviewers, and conflicts regarding relevance of a record were resolved by a third reviewer.

**Table 1.** Definitions and types of Master protocol trial designs.

| Terminology | Definition |
| --- | --- |
| Master protocol | Trial design that evaluates multiple diseases, multiple treatments, or both under a single trial infrastructure and protocol [1]. |
| Basket trial | Evaluates one targeted therapy on multiple diseases or multiple disease subtypes. |
| Umbrella trials | Evaluates multiple targeted therapies for different subgroups of a single disease [11]. |
| Platform trial | Evaluates multiple experimental treatments in a single disease in a perpetual manner [1]. |
| Complex design | A trial design that combine features of any two or more master protocol trial designs (for instance both umbrella and basket features). |

**Table 2.** Trial characteristics of identified Master protocols.

| Variable | Basket (N=31) |  | Umbrella (N=29) |  | Platform (N=36) |  | Complex designs (N=6) |  | Total (N=102) |
| --- | --- | --- | --- | --- | --- | --- | --- | --- | --- |
|  | LMIC incl. | LMIC excl. | LMIC incl. | LMIC excl. | LMIC incl. | LMIC excl. | LMIC incl. | LMIC excl. |  |
| **Disease setting** |  |  |  |  |  |  |  |  |  |
| Oncology | 16 (51.6%) | 1 (3.2%) | 23 (79.3%) | 2 (6.9%) | 7 (19.4%) | 1 (2.8%) | 5 (83.3%) | 1 (16.7%) | 56 (54.9%) |
| Non-oncology | 12 (38.7%) | 2 (6.5%) | 4 (13.8%) | 0 (0.0%) | 10 (27.8%) | 18 (50.0%) | 0 (0.0%) | 0 (0.0%) | 46 (45.1%) |
| **Trial phase** |  |  |  |  |  |  |  |  |  |
| Phase I | 2 (6.5%) | 0 (0.0%) | 1 (3.4%) | 0 (0.0%) | 1 (2.8%) | 0 (0.0%) | 0 (0.0%) | 0 (0.0%) | 4 (3.9%) |
| Phase II | 14 (45.2%) | 1 (3.2%) | 14 (48.3%) | 1 (3.4%) | 5 (13.9%) | 6 (16.7%) | 4 (66.7%) | 1 (16.7%) | 46 (45.1%) |
| Phase I/II | 3 (9.7%) | 0 (0.0%) | 11 (37.9%) | 1 (3.4%) | 3 (8.3%) | 2 (5.6%) | 0 (0.0%) | 0 (0.0%) | 20 (19.6%) |
| Phase III | 7 (22.6%) | 1 (3.2%) | 0 (0.0%) | 0 (0.0%) | 3 (8.3%) | 6 (16.7%) | 0 (0.0%) | 0 (0.0%) | 17 (16.7%) |
| Phase II/III | 0 (0.0%) | 0 (0.0%) | 0 (0.0%) | 0 (0.0%) | 3 (8.3%) | 3 (8.3%) | 0 (0.0%) | 0 (0.0%) | 6 (5.9%) |
| Phase IV | 0 (0.0%) | 0 (0.0%) | 1 (3.4%) | 0 (0.0%) | 1 (2.8%) | 2 (5.6%) | 0 (0.0%) | 0 (0.0%) | 4 (3.9%) |
| Unspecified | 2 (6.5%) | 1 (3.2%) | 0 (0.0%) | 0 (0.0%) | 1 (2.8%) | 0 (0.0%) | 1 (16.7%) | 0 (0.0%) | 5 (4.9%) |
| **Study sponsor** |  |  |  |  |  |  |  |  |  |
| Industry-funded | 24 (77.4%) | 1 (3.2%) | 14 (48.3%) | 2 (6.9%) | 8 (22.2%) | 9 (25.0%) | 3 (50.0%) | 1 (16.7%) | 60 (58.8%) |
| Government funded | 4 (12.9%) | 2 (6.5%) | 13 (44.8%) | 0 (0.0%) | 9 (25.0%) | 10 (27.8%) | 2 (33.3%) | 0 (0.0%) | 42 (41.2%) |
| **Sponsor category** |  |  |  |  |  |  |  |  |  |
| Pharma | 23 (74.2%) | 1 (3.2%) | 13 (44.8%) | 2 (6.9%) | 8 (22.2%) | 5 (13.9%) | 3 (50.0%) | 1 (16.7%) | 56 (55.0%) |
| Academia | 1 (3.2%) | 1 (3.2%) | 8 (27.6%) | 0 (0.0%) | 6 (16.7%) | 8 (22.2%) | 1 (16.7%) | 0 (0.0%) | 26 (25.5%) |
| Govt. research institutes | 1 (3.2%) | 0 (0.0%) | 1 (3.4%) | 0 (0.0%) | 2 (5.6%) | 2 (5.6%) | 0 (0.0%) | 0 (0.0%) | 6 (5.9%) |
| Hospitals | 2 (6.5%) | 0 (0.0%) | 4 (13.8%) | 0 (0.0%) | 1 (2.8%) | 0 (0.0%) | 1 (16.7%) | 0 (0.0%) | 8 (7.8%) |
| Non-profit organisation | 0 (0.0%) | 0 (0.0%) | 1 (3.4%) | 0 (0.0%) | 0 (0.0%) | 2 (5.6%) | 0 (0.0%) | 0 (0.0%) | 3 (2.9%) |
| Investigator sponsored | 1 (3.2%) | 0 (0.0%) | 0 (0.0%) | 0 (0.0%) | 0 (0.0%) | 2 (5.6%) | 0 (0.0%) | 0 (0.0%) | 3 (2.9%) |
| **Completion status** |  |  |  |  |  |  |  |  |  |
| Completed | 2 (6.5%) | 0 (0.0%) | 1 (3.4%) | 0 (0.0%) | 2 (5.6%) | 6 (16.7%) | 0 (0.0%) | 0 (0.0%) | 11 (10.8%) |
| Not started | 1 (3.2%) | 1 (3.2%) | 4 (13.8%) | 0 (0.0%) | 1 (2.8%) | 0 (0.0%) | 0 (0.0%) | 0 (0.0%) | 7 (6.8%) |
| Ongoing | 18 (58.1%) | 2 (6.5%) | 21 (72.4%) | 2 (6.9%) | 12 (33.3%) | 10 (27.8%) | 4 (66.7%) | 0 (0.0%) | 69 (67.7%) |
| Suspended/terminated | 4 (12.9%) | 0 (0.0%) | 1 (3.4%) | 0 (0.0%) | 2 (5.6%) | 3 (8.3%) | 0 (0.0%) | 1 (16.7%) | 11 (10.8%) |
| Unknown | 3 (9.7%) | 0 (0.0%) | 0 (0.0%) | 0 (0.0%) | 0 (0.0%) | 0 (0.0%) | 1 (16.7%) | 0 (0.0%) | 7 (6.8%) |
| **Study population** |  |  |  |  |  |  |  |  |  |
| Adolescents | 1 (3.2%) | 0 (0.0%) | 0 (0.0%) | 0 (0.0%) | 0 (0.0%) | 0 (0.0%) | 0 (0.0%) | 0 (0.0%) | 1 (1.0%) |

<sup>a</sup>Complex designs refer to trial designs that do not neatly fit the working definitions adopted in Table 1; LMIC excl. refers to the category of trials run exclusively in LMIC countries; LMIC incl. refers to the category of trials run in at least one LMIC country.

Table 2 continued...: Trial characteristics of identified Master protocols
| Variable | Basket (N=31) |  | Umbrella (N=29) |  | Platform (N=36) |  | Complex designs (N=6) |  | Total (N=102) |
| --- | --- | --- | --- | --- | --- | --- | --- | --- | --- |
|  | LMIC incl. | LMIC excl. | LMIC incl. | LMIC excl. | LMIC incl. | LMIC excl. | LMIC incl. | LMIC excl. |  |
| Adults | 20 (64.5%) | 3 (9.7%) | 27 (93.1%) | 2 (6.9%) | 15 (41.7%) | 16 (44.4%) | 4 (66.7%) | 1 (16.7%) | 88 (86.3%) |
| Adolescents Adults | 1 (3.2%) | 0 (0.0%) | 0 (0.0%) | 0 (0.0%) | 0 (0.0%) | 3 (8.3%) | 0 (0.0%) | 0 (0.0%) | 4 (3.9%) |
| Children Adolescents | 2 (6.5%) | 0 (0.0%) | 0 (0.0%) | 0 (0.0%) | 1 (2.8%) | 0 (0.0%) | 0 (0.0%) | 0 (0.0%) | 3 (2.9%) |
| All | 4 (12.9%) | 0 (0.0%) | 0 (0.0%) | 0 (0.0%) | 1 (2.8%) | 0 (0.0%) | 1 (16.7%) | 0 (0.0%) | 6 (5.9%) |
| **Eligible Sex** |  |  |  |  |  |  |  |  |  |
| Female | 1 (3.2%) | 0 (0.0%) | 9 (31.0%) | 0 (0.0%) | 3 (8.3%) | 0 (0.0%) | 0 (0.0%) | 0 (0.0%) | 13 (12.7%) |
| Both | 27 (87.1%) | 3 (9.7%) | 18 (62.1%) | 2 (6.9%) | 14 (38.9%) | 19 (52.8%) | 5 (83.3%) | 1 (16.7%) | 89 (87.3%) |
| **Type of interventions** |  |  |  |  |  |  |  |  |  |
| Pharmaceutical | 26 (83.9%) | 2 (6.5%) | 26 (89.7%) | 2 (6.9%) | 16 (44.4%) | 19 (52.8%) | 5 (83.3%) | 1 (16.7%) | 97 (95.1%) |
| Non-pharmaceutical | 2 (6.5%) | 1 (3.2%) | 0 (0.0%) | 0 (0.0%) | 1 (2.8%) | 0 (0.0%) | 0 (0.0%) | 0 (0.0%) | 4 (3.9%) |
| Both | 0 (0.0%) | 0 (0.0%) | 1 (3.4%) | 0 (0.0%) | 0 (0.0%) | 0 (0.0%) | 0 (0.0%) | 0 (0.0%) | 1 (1.0%) |
| **Study published** |  |  |  |  |  |  |  |  |  |
| Yes | 4 (13.0%) | 0 (0.0%) | 4 (13.8%) | 0 (0.0%) | 6 (16.7%) | 9 (25.0%) | 1 (16.7%) | 1 (16.7%) | 25 (24.5%) |
| No | 24 (77.4%) | 3 (9.7%) | 23 (79.2%) | 2 (6.8%) | 11 (30.6%) | 11 (27.7%) | 4 (66.7%) | 0 (0.0%) | 77 (75.5%) |
| **Published material** |  |  |  |  |  |  |  |  |  |
| Study protocol | 1 (3.2%) | 0 (0.0%) | 0 (0.0%) | 0 (0.0%) | 0 (0.0%) | 2 (5.6%) | 1 (16.7%) | 0 (0.0%) | 4 (3.9%) |
| Study design | 0 (0.0%) | 0 (0.0%) | 2 (6.9%) | 0 (0.0%) | 1 (2.8%) | 0 (0.0%) | 0 (0.0%) | 0 (0.0%) | 3 (2.9%) |
| Results | 3 (9.6%) | 0 (0.0%) | 1 (3.4%) | 0 (0.0%) | 4 (11.1%) | 5 (13.9%) | 0 (0.0%) | 0 (0.0%) | 13 (12.8%) |
| Study design Results | 0 (0.0%) | 0 (0.0%) | 1 (3.4%) | 0 (0.0%) | 1 (2.8%) | 2 (5.6%) | 0 (0.0%) | 1 (16.7%) | 5 (4.9%) |
| None | 24 (77.4%) | 3 (9.7%) | 23 (79.3%) | 2 (6.9%) | 11 (30.6%) | 10 (27.8%) | 4 (66.7%) | 0 (0.0%) | 77 (75.5%) |
| **Trial documents** |  |  |  |  |  |  |  |  |  |
| Study protocol | 1 (3.2%) | 0 (0.0%) | 0 (0.0%) | 0 (0.0%) | 2 (5.6%) | 3 (8.3%) | 0 (16.7%) | 0 (0.0%) | 7 (6.9%) |
| Study protocol SAP | 2 (6.4%) | 0 (0.0%) | 0 (0.0%) | 0 (0.0%) | 1 (2.8%) | 1 (2.8%) | 0 (0.0%) | 0 (0.0%) | 4 (3.9%) |
| None | 25 (80.6%) | 3 (9.7%) | 27 (93.1%) | 2 (6.9%) | 14 (38.9%) | 15 (41.7%) | 4 (66.7%) | 1 (16.7%) | 91 (89.2%) |

<sup>a</sup>Complex designs refer to trial designs that do not neatly fit the working definitions adopted in Table 1; LMIC excl. refers to the category of trials run exclusively in LMIC countries; LMIC incl. refers to the category of trials run in at least one LMIC country.

### 2.3 Data extraction and synthesis

Each individual trial record was independently extracted for information by two reviewers. We extracted information on trial identifiers (trial ID and acronyms), master protocol type, trial phase, disease area, the number of modules (subtrials in an umbrella design; disease subgroups in a basket trial; arms including the control in a platform trial), the sample size, randomisation, error rate control, the primary outcome, any adaptive features, the analysis approach (Bayesian or frequentist), trial sponsor, countries covered, recruitment and completion status, amongst other considerations. A listing of all information extracted from the trial registry is available in the supplementary material. The data were then synthesised into a single dataset, from which the descriptive analysis was undertaken.

## 3 Results

We retrieved a total of 10,281 trial records across the various registries from our initial search. After removal of duplicates, and records not meeting eligibility criteria, 189 trial records of 102 unique master protocol trials were included in our study. We found 31 basket trials, 29 umbrella trials, 36 platform trials and 6 complex master protocol designs conducted in 54 countries. A complete list of all trial records is available in the supplementary material Table 4.

### 3.1 Trends of master protocols in LMICs

Master protocols have been gaining popularity in LMICs, particularly over the last five years (2019 onwards) (Figure 1). Notably, the majority of the trials reported in this review (93/102, 91.2%) launched after the onset of the COVID-19 pandemic. The earliest master protocol reported in our study was the DIAN-TU [6] platform trial in Alzheimers disease that started in 2012 and ran across 13 countries, 3 of which were LMICs (Argentina, Colombia and Mexico). The earliest master protocol ran exclusively in LMIC sites was TAC [29] in 2015, an umbrella trial in Hepatitis C Virus-infected Patients in West and Central Africa.

**Fig 1.**
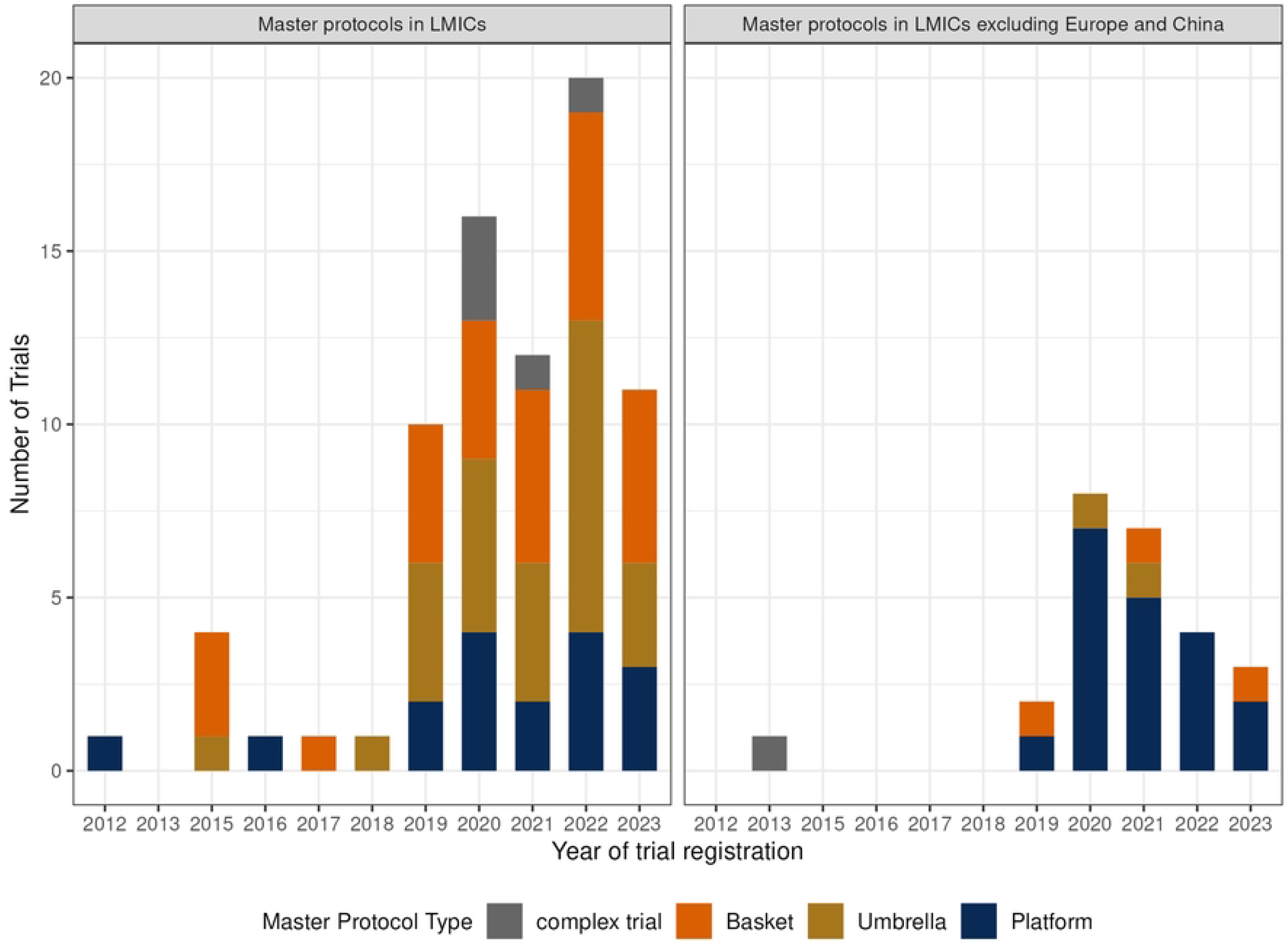
Trends of master protocols in LMICs over time. Our results show that most master protocols in LMICs are run in China (n=52), European LMICs like Turkey (n=18) and Russia (n=16), and upper middle-income countries such as Brazil (n=22), Argentina (n=16), Mexico (n=14), South Africa (n=12), India (n=7), and Colombia (n=6). In Figure 2, we observe that while the majority of trials were multi-country studies, the proportion of LMICs was often less than 30%, figures that were lower when not considering China and LMICs in Europe. For instance, nearly half (48/102, 47.1%) of the trials did not have an LMIC site outside of China and Europe. Excluding European LMICs and China, there are far fewer master protocol trials ongoing in other LMIC regions. The majority of these were platform trials (19/102, 18.6%) followed by basket (3/102, 2.9%), and then umbrella trials (2/102, 1.9%).

**Fig 2.**
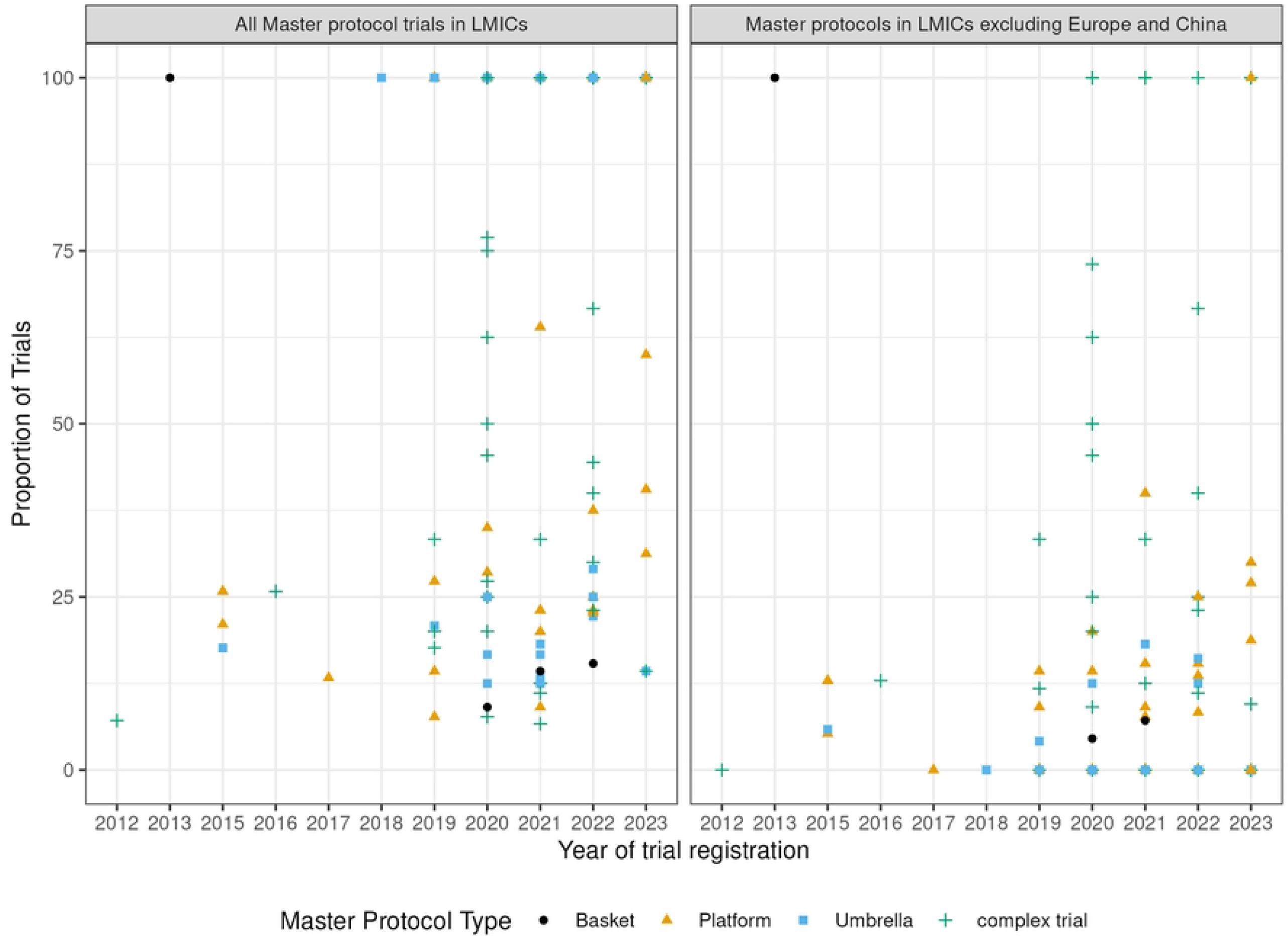
Proportion of LMIC country representation in master protocols. Each point on the graph represents a unique trial included in our review. The proportions are derived as the number of LMIC countries in a specific trial over total number of countries where the trial is run globally

### 3.2 Characteristics of master protocols in LMICs

We present a summary of the master protocol trial characteristics in Table 3. Oncology was the most common therapeutic area (54.9%, 56/102) where master protocols are conducted/ongoing, mainly featuring umbrella and basket trials. There was an increase in the uptake of master protocols in both oncology and non-oncology areas over time. Between 2012 and 2018, we identified 2.9% of master protocols (3/102) in oncology and 3.9% (4/102) in non-oncology. From 2019 to 2023, this increased significantly, with 54.9% (56/102) in oncology and 45.1% (46/102) in non-oncology. Thus, the aforementioned trend in the rise of master protocols in LMICs was consistently reflected across both oncology and non-oncology fields. Notable, the most common disease area used in non-oncology was COVID-19 and other respiratory diseases. Platform trials were the most common master protocol design used in non-oncology (28/46, 60.9%), followed by basket trials (14/46, 30.4%).

**Table 3.** Statistical characteristics of identified Master protocols.

| Variable | Basket (N=31) |  | Umbrella (N=29) |  | Platform (N=36) |  | Complex designs (N=6) |  | Total (N=102) |
| --- | --- | --- | --- | --- | --- | --- | --- | --- | --- |
|  | LMIC incl. | LMIC excl. | LMIC incl. | LMIC excl. | LMIC incl. | LMIC excl. | LMIC incl. | LMIC excl. |  |
| **Randomised<sup>c</sup>** |  |  |  |  |  |  |  |  |  |
| Yes | 9 (29.0%) | 2 (6.5%) | 13 (44.8%) | 1 (3.4%) | 14 (38.9%) | 19 (52.8%) | 1 (16.7%) | 0 (0.0%) | 59 (57.8%) |
| No | 19 (61.3%) | 1 (3.2%) | 14 (48.3%) | 1 (3.4%) | 3 (8.3%) | 0 (0.0%) | 4 (66.7%) | 1 (16.7%) | 43 (42.2%) |
| **Adaptive design<sup>c</sup>** |  |  |  |  |  |  |  |  |  |
| Yes | 8 (25.8%) | 0 (0.0%) | 17 (58.6%) | 1 (3.4%) | 17 (47.2%) | 19 (52.8%) | 1 (16.7%) | 1 (16.7%) | 64 (62.7%) |
| No | 14 (45.2%) | 2 (6.5%) | 6 (20.7%) | 1 (3.4%) | 0 (0.0%) | 0 (0.0%) | 1 (16.7%) | 0 (0.0%) | 24 (23.5%) |
| Unclear | 6 (19.4%) | 1 (3.2%) | 4 (13.8%) | 0 (0.0%) | 0 (0.0%) | 0 (0.0%) | 3 (50.0%) | 0 (0.0%) | 14 (13.8%) |
| **Adaptive design type<sup>c</sup>** |  |  |  |  |  |  |  |  |  |
| Adding and dropping arms | 1 (3.1%) | 0 (0.0%) | 11 (36.7%) | 1 (3.3%) | 17 (47.2%) | 19 (52.8%) | 0 (0.0%) | 0 (0.0%) | 46 (44.2%) |
| Group Sequential | 3 (9.4%) | 0 (0.0%) | 0 (0.0%) | 0 (0.0%) | 0 (0.0%) | 3 (8.3%) | 1 (16.7%) | 0 (0.0%) | 7 (6.7%) |
| Adaptive randomisation | 0 (0.0%) | 0 (0.0%) | 0 (0.0%) | 0 (0.0%) | 1 (2.8%) | 2 (5.6%) | 0 (0.0%) | 0 (0.0%) | 3 (2.9%) |
| Adaptive treatment switching | 1 (3.1%) | 0 (0.0%) | 0 (0.0%) | 0 (0.0%) | 0 (0.0%) | 0 (0.0%) | 0 (0.0%) | 0 (0.0%) | 1 (1.0%) |
| Sample size re-estimation | 0 (0.0%) | 0 (0.0%) | 0 (0.0%) | 0 (0.0%) | 0 (0.0%) | 1 (2.8%) | 0 (0.0%) | 0 (0.0%) | 1 (1.0%) |
| Seamless design | 3 (9.4%) | 0 (0.0%) | 11 (36.7%) | 1 (3.3%) | 5 (13.9%) | 4 (11.1%) | 0 (0.0%) | 0 (0.0%) | 24 (23.1%) |
| Dose adaptations | 3 (9.4%) | 0 (0.0%) | 2 (6.7%) | 0 (0.0%) | 2 (5.6%) | 1 (2.8%) | 0 (0.0%) | 0 (0.0%) | 8 (7.7%) |
| Efficacy/futility stopping | 0 (0.0%) | 0 (0.0%) | 0 (0.0%) | 0 (0.0%) | 0 (0.0%) | 1 (2.8%) | 0 (0.0%) | 1 (16.7%) | 2 (2.0%) |
| Unclear/None | 0 (0.0%) | 0 (0.0%) | 0 (0.0%) | 0 (0.0%) | 0 (0.0%) | 1 (2.8%) | 0 (0.0%) | 1 (16.7%) | 38 (37.3%) |
| **Bayesian/Freq. design<sup>c</sup>** |  |  |  |  |  |  |  |  |  |
| Bayesian | 1 (3.2%) | 0 (0.0%) | 3 (10.3%) | 0 (0.0%) | 3 (8.4%) | 7 (19.5%) | 0 (0.0%) | 0 (0.0%) | 14 (13.7%) |
| Frequentist | 24 (77.5%) | 3 (9.7%) | 21 (72.4%) | 2 (6.9%) | 11 (30.6%) | 9 (25.0%) | 4 (66.7%) | 1 (16.7%) | 77 (75.5%) |
| Unclear/unspecified | 3 (9.7%) | 0 (0.0%) | 3 (10.3%) | 0 (0.0%) | 3 (8.3%) | 1 (2.8%) | 1 (16.7%) | 0 (0.0%) | 11 (10.8%) |
| **Pooled analysis<sup>c</sup>** |  |  |  |  |  |  |  |  |  |
| Yes | 3 (9.7%) | 0 (0.0%) | 2 (6.9%) | 0 (0.0%) | 0 (0.0%) | 1 (2.8%) | 3 (50.0%) | 0 (0.0%) | 9 (8.8%) |
| No | 8 (25.8%) | 1 (3.2%) | 11 (37.9%) | 0 (0.0%) | 8 (22.2%) | 4 (11.1%) | 1 (16.7%) | 1 (16.7%) | 34 (33.3%) |
| Unclear/unspecified | 17 (54.8%) | 2 (6.5%) | 14 (48.3%) | 2 (6.9%) | 9 (25.0%) | 13 (38.9%) | 1 (16.7%) | 0 (0.0%) | 59 (57.9%) |
| **Common control<sup>c</sup>** |  |  |  |  |  |  |  |  |  |
| Yes | 9 (29.0%) | 0 (0.0%) | 10 (34.5%) | 1 (3.4%) | 12 (33.3%) | 18 (50.0%) | 0 (0.0%) | 0 (0.0%) | 50 (49.0%) |
| No | 19 (61.3%) | 3 (9.7%) | 17 (58.6%) | 1 (3.4%) | 5 (13.9%) | 1 (2.8%) | 5 (83.3%) | 1 (16.7%) | 52 (51.0%) |

<sup>a</sup>Complex designs refer to trial designs that do not neatly fit the working definitions adopted in Table 1; LMIC excl. refers to the category of trials run exclusively in LMIC countries; LMIC incl. refers to the category of trials run in at least one LMIC country.

Most of the master protocols (67.7%) are still ongoing, with just 11/102 studies (10.8%) having been completed. A small proportion of trials (10.8%) have either been withdrawn, suspended or terminated, and 7 studies (6.8%) registered on a trial database as of our last search (September 2023), were yet to start. In terms of trial phases, the majority of master protocols were early phase studies (70%) (i.e., phase I or phase II studies), compared to late phase studies. While umbrella, basket and platform designs were equally common in early-phase studies, late-phase studies were often dominated by platform trials.

Our findings further show that the master protocols were commonly industry-sponsored studies (58.8%) investigating pharmaceutical interventions (95.1%) and conducted in adult populations (*≥* 18 Years old; 86.3%), enrolling both sexes (87.3%). Trial documentation such as study protocol and statistical analysis plans (SAP) were mostly unavailable (89.2%), and only a few studies had been published (75.5%), due to the smaller proportion of completed studies.

### 3.3 Design considerations of the master protocols

We present the trial design properties for the master protocols in Table 3. Among the 102 studies, we observed common use of randomisation (57.8%), adaptive design features (62.7%), and a frequentist approach to trial design (75.5%).

The most common adaptive features were adding and dropping of arms (23.5%), often a feature of platform trials, and seamless designs (22.5%) i.e., phase I/II and phase II/III studies, followed by group sequential designs (7.8%) and dose adaptations (7.8%). These adaptive design features including adaptive randomisation and sample-size re-estimation were commonly present in master protocols. Seamless designs were commonly present in umbrella and platform trials, and rarely in basket studies, partly attributed to the several treatments under investigation in umbrella and platform designs. Bayesian approaches to trial design was less common overall (13.7%) but were more commonly used in platform trials (10/36, 27.8%).

However, in more than one-third of trial records (37.3%) it was unclear whether any adaptive features were present, largely attributed to the widespread unavailability of relevant trial documentation such as SAPs and trial protocols in trial databases.

Complex designs had on average higher number of novel treatments under investigation (median = 10), the highest number of subtrials/modules (median = 10.5), larger sample sizes (median= 596), and longer trial duration (median = 5 months) compared to the conventional master protocols like umbrella, platform, and basket trials(Figure 3).

**Fig 3.**
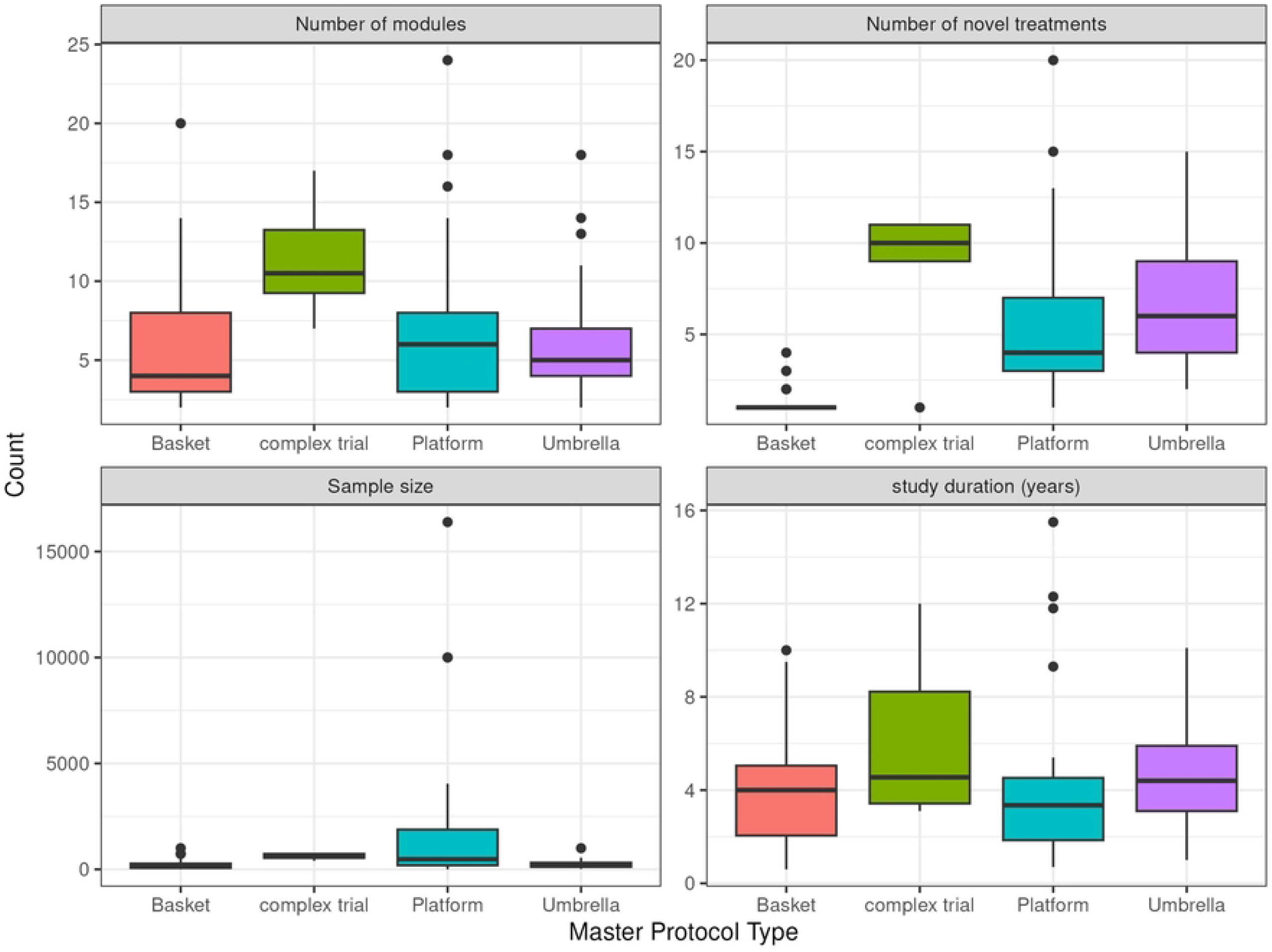
Summary of novel treatments, modules, sample size and study duration of the Master Protocols in LMICs. In terms of the conventional master protocols, platform trials, on average, had the highest number of subtrials/modules (median = 6) and recruited the largest sample size (median = 477) when compared to umbrella and basket trials. However, they had a shorter study duration (median = 3.4 years) and included fewer novel treatments (median = 4) compared to umbrella trials. This is unsurprising of platform trials given their varied use across different therapeutic areas, and their common use of adaptive design.

### 3.4 Geographical distribution of Master protocols in LMICs

We present the geographical distribution of the master protocols in Figure 4 and supplementary Table 1. Studies were mostly multi-country trials (65.7%), with a greater representation from China, Brazil, and Russia. There were only 5 studies conducted exclusively in LMIC outside China and Europe, and these were three platform studies in COVID-19, a single platform study in Tuberculosis and one multi-basket study in HER2/EGFR solid tumours. Of these, 4/5 studies were conducted exclusively in Africa, except for one study in COVID-19(Table 4). One trial in HER2/EGFR solid tumours was terminated, while the rest are currently ongoing. Thus, at the time of writing, no trial evaluating targeted therapies under a master protocol framework has been designed, run and completed in Africa to the best of our knowledge.

**Fig 4.**
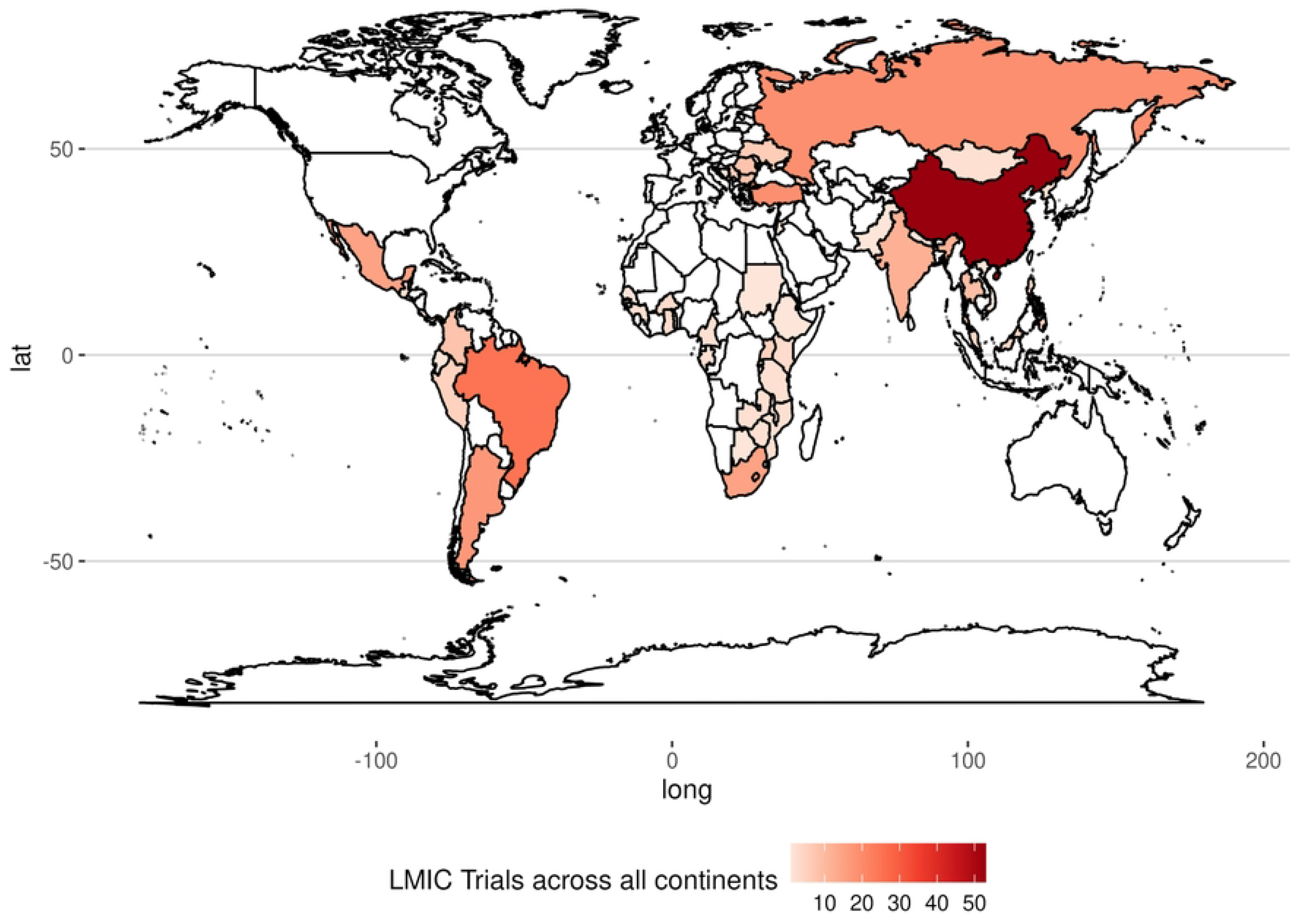
Geographical representation of master protocols in LMICs. The map is generated from data in our review. Dense colors represent a higher number of master protocols in those regions

**Table 4.** Highlighted examples of ongoing or completed Master protocol trials in LMICs.

| Trial ID/registration | Trial design | Disease setting | Description |
| --- | --- | --- | --- |
| EUCTR2020-004457-76 | Basket trial | Crohn Disease, Ulcerative Colitis and Juvenile Psoriatic Arthritis | A Phase 3, Multicenter, Open-label, Basket, Long-term Extension Study of Ustekinumab in Pediatric Clinical Study Participants aged 2 to 18 years across 13 countries. |
| ACTG A5288 | Umbrella trial | HIV | An open-label phase IV study of HIV-1 infected participants with triple-class experience or resistance to nucleoside reverse transcriptase inhibitors (NRTIs), non-NRTIs (NNRTIs), and protease inhibitors (PIs) and who were failing their current regimen. Patients were assigned to one of four Cohorts (A, B, C or D) based on their screening genotype results and antiretroviral (ARV) history where novel agents are investigated in each cohort |
| ANTICOV<br>PACTR202006537901307 | Platform trial | COVID-19 | An open-label, randomised, adaptive platform trial testing the safety and efficacy of treatments in mild-to-moderate COVID-19 patients. The trial aimed to identify early treatments that could prevent progression of COVID-19 to severe disease and potentially limit transmission. |
| NCT04589845 | Complex trial | Solid tumors | A Phase II, global, multicenter, open-label, multi-cohort study to evaluate the safety and efficacy of targeted therapies or immunotherapy in participants with unresectable, locally advanced or metastatic solid tumors that harbor specific oncogenic genomic alterations or who are tumor mutational burden (TMB)-high as identified by a validated next-generation sequencing (NGS) assay. |

Regionally, most studies were conducted in Asia, South-America and Europe (82.3%), with a few studies spread across Africa, South-Africa accounting for a greater majority. The median number of countries per trial was 7 (IQR: 1 to 13) overall, and the median number of LMICs per trial was 1. Excluding China and European LMIC representation, the median number of LMICs per trial was only 1.

## Discussion

Although master protocol clinical trials efficiently answer multiple research questions, low and middle-income countries still remain underrepresented in global research of this type. We reviewed trial registries to understand the extent of implementation of master protocols, and found increasing implementation of master protocols in LMICs, but that is largely dominated by China and middle-income countries in Europe.

The under-representation of LMICs in master protocol trials is concerning, considering the potential benefits that these designs could offer in these setting where resources for conducting traditional large scale trials are limited. Master protocols could provide an efficient approach to study multiple treatments and indications (including rare diseases), thus optimising resources. The limited use of master protocols outside of China and European LMICs highlights the potential existence of unique challenges to the implementation of master protocols in LMICs. In these settings, limited infrastructure and financial resources for managing complex designs, ethical and regulatory system obstacles, operational barriers have been reported as critical barriers to conducting trials in developing countries [30]. Besides, the limited penetration of precision medicine and related technology in most LMICs [31], and lack of expertise to design and implement these designs in lower-income countries have further limited their use.

This review, similarly to recent reviews [13, 32], reports a higher prevalence of master protocols in oncology and early phase trial settings. However, of interest to note is the increasing utilisation of master protocols in non-oncology areas. For LMICs, this signals new opportunities to leverage the benefits of master protocols to address important treatment-related questions in infectious and non-communicable diseases that are leading public health concerns in specific LMIC regions such as Africa. Diseases such as HIV, Ebola, and COVID-19 have welcomed the use of umbrella and platform trials, yet still there are opportunities for indications such as Tuberculosis, neglected tropical diseases to benefit from these innovative designs to accelerate the pace of evidence generation for the introduction of life-saving therapies to meet the needs of patients in resource poor settings.

The increased awareness and opportunities afforded by master protocols in both oncology and non-oncology settings, however, presents both opportunities and challenges for LMICs. To ensure LMICs fully capitalize on this evolving trend, it is imperative that several key considerations are addressed. Firstly, there is an urgent need for capacity-building initiatives for stakeholders including regulators, researchers, patient advocacy groups. This includes initiatives aimed at strengthening clinical trial infrastructure in LMICs, training researchers in master protocols design and conduct, improving regulatory frameworks, and enhancing data management capabilities. Without such foundational elements, LMICs will continue to lag behind in the global master protocol trials landscape, and in implementing their own regionally or locally relevant studies.

For several trials, the principal investigators and even statistical expertise was mostly domiciled in HICs. For example, the statistical expertise behind the design of some of the largest platform trials in Africa such as the EBOLA platform trial [33] and ANTICOV trial in COVID-19 [34], is all domiciled in HICs. Such is the case for many other master protocols in our review. However, this is unsurprising given the global inequities in precision medicine research [35]. This dependency on external expertise for several master protocols raises critical questions about readiness of LMICs to spearhead their own research in times such as the recent pandemic requiring the launch of efficient designs to investigate treatments faster and more efficiently.

Though commonly multi-country studies, the skewed nature of existing collaborations in recent and completed master protocols - involving high income countries and select LMICs, often where clinical trial infrastructure is well established, highlight the need for further collaborative efforts between high-income countries (HICs) and LMICs. Partnerships between HICs and LMICs can facilitate knowledge transfer, promote capacity-building, and provide financial and technical support to overcome the barriers that LMICs currently face [31, 36]. For example, joint research initiatives or public-private partnerships could pool resources and expertise to launch master protocol trials targeting health challenges specific to LMICs. In this way, LMICs could not only benefit from cutting-edge scientific methodologies but also contribute to global health innovation.

Additionally, regulatory harmonization between HICs and LMICs is vital to ensure that master protocols can be efficiently implemented across different regions. Given that master protocols are inherently more complex than traditional trial designs, regulatory bodies in LMICs need support in adapting their review processes to accommodate the flexibility of these trials. Even in HICs, implementers of master protocols experienced hurdles with regulatory agencies, including varying levels of acceptance of master protocol trial submissions (particularly in the confirmatory setting) and inconsistent regulatory feedback [32]. We anticipate the inexperience of agencies in LMICs can pose a considerable challenge to adoption of these trials, being in favour of less complex proposals. As such, regional collaborations or capacity-building initiatives within regulatory agencies could improve oversight while maintaining high standards of safety and efficacy.

In conclusion, while the current landscape of master protocols shows ongoing progress, there is significant potential for broader adoption in LMICs, particularly in non-oncology settings. We argue, that there are considerable benefits to be gained in drug development through wider implementation of master protocols in LMIC settings. Strategic efforts aimed at enhancing clinical trial capacity, fostering cross-border collaboration, and harmonizing regulatory standards will be crucial in realizing the full potential of master protocols in LMICs. Such efforts are necessary to ensure that LMICs are not left behind in the global shift towards more innovative and efficient trial designs, ultimately improving the speed at which new treatments reach patients in these regions.

## Funding

JMSW is funded by a NIHR Research Professorship (NIHR301614).

## Data availability statement

All data relevant to this study has been included in the article or available in the supplementary material.

## Notes

### Competing Interest Statement

The authors have declared no competing interest.

### Author Declarations

This study utilised publicly available data, that is available in our manuscript or the supplementary material.

